# Low-frequency Oscillations in Postural Sway Reflect Sensory Reweighting but Become Decoupled from Postural Output as Huntington’s Disease Progresses

**DOI:** 10.64898/2026.05.19.26353619

**Authors:** Marina Meyer Vega, Tyler Wadlington, Kyle T Gunning, Alexander J Lytle, Juan Pablo Murcia, Anthony J Percuoco, Niyati Baweja, Daniel J Goble, Paul E Gilbert, Jody Corey-Bloom, Harsimran S Baweja

**Affiliations:** School of Kinesiology, Auburn University, Auburn, AL, USA; School of Physical Therapy, San Diego State University, San Diego, CA, USA; School of Health Science, Oakland University, Rochester, MI, USA; Department of Psychology, College of Science, San Diego State University, San Diego, CA, USA; Department of Neuroscience, University of California at San Diego, San Diego, CA, USA

**Author notes:** **Corresponding author:** Harsimran S. Baweja P.T., Ph.D., Director of the Auburn University Physical Therapy Program, Director of SensoriMotor and Rehabilitation Technology Neuroscience Lab, School of Kinesiology, Auburn University, 301 Wire Road, Auburn University, Auburn, AL 36849.

**Keywords:** Huntington’s disease, postural sway, low-frequency oscillations, sensory reweighting, premanifest HD

## Abstract

**Background:** Huntington’s disease (HD) causes progressive postural control deficits, but how sensory reweighting mechanisms degrade across disease stages remains poorly understood.

**Objective:** To determine whether objective markers of postural sway track disease severity and altered sensory reweighting across the HD spectrum.

**Methods:** Ninety-seven adults (46±14 yrs) were categorized into four groups: 29 with HD, 27 pre-manifest (PM), 28 not at risk (AR-), and 13 age-matched healthy controls (HC). Participants performed three trials of quiet standing with eyes open and eyes closed on a force plate.

**Results:** Manifest HD individuals exhibited greater AP, ML, and total COP sway displacement compared with the PM, AR-, and HC groups. HD and PM groups demonstrated greater instability with eyes closed. COP wavelet power was concentrated below 1 Hz across all groups. The eyes-open to eyes-closed change in 0-1 Hz power predicted total COP sway in HC (68%), AR- (45%), and PM (46%), but this relation was substantially weaker in HD.

**Conclusions:** Progressive weakening of oscillatory-sway coupling distinguishes manifest HD from premanifest stages. PM individuals demonstrate early sensory reweighting deficits that manifest only when vision is removed, while HD individuals show decoupled oscillatory activity that fails to support stable postural regulation. This progressive decoupling may serve as a candidate marker of disease conversion prior to overt motor diagnosis.

## Introduction

Huntington’s disease (HD) is a progressive neurodegenerative disorder caused by a CAG repeat expansion in the HTT gene, characterized clinically by motor impairment and cognitive decline.^1–4^ Postural control is a critical clinical feature of HD because the basal ganglia integrate proprioceptive, visual, and vestibular information to maintain upright stance.^5,6^ As striatal neurons are lost, this integration breaks down, producing altered sensorimotor processing, impaired muscle tone regulation, difficulty with fine motor tasks, and chorea.^6–8^

Balance impairment is an early feature of manifest HD that may begin in the premanifest stage and worsen with disease progression.^9–12^ Assessing balance under degraded visual or proprioceptive input could identify the transition point at which premanifest individuals (PM) develop clinically manifest disease.^11^ However, studies mapping postural control across the HD spectrum, especially using family members who are at risk negative (AR-) as controls, remain limited. AR- individuals provide a unique genetic control: they share the environmental and familial background of HD families but lack CAG expansion, isolating HD-specific deficits.^13^

Traditional clinical balance measures rely on task completion time or subjective scoring, such as the Berg Balance Scale and the Timed Up and Go.^14,15^ Although linear center of pressure (COP) measures objectively quantify the magnitude of postural sway, they reduce a rich physiological signal to total displacement (cm), ignoring its time-frequency structure.^16^ As a result, they say little about which sensory systems contribute to balance regulation, or how these contributions shift under specific task demands.^16,17^

Time-frequency analyses of the COP signal can resolve task-, age-, and disease-related differences in postural control, and have been proposed as markers of postural control decline.^16,18,19^ Much of the relevant oscillatory information in the COP signal lies between 0 and 4 Hz,^19,20^ and within this range, modulation of low-frequency oscillations is thought to reflect sensory reweighting among visual, vestibular, and proprioceptive inputs.^16,21,22^

However, whether low-frequency patterns differ across AR-, PM, and manifest HD remains unknown, limiting our ability to determine whether changes in the time-frequency structure of postural sway constitute disease-specific markers of HD conversion. Therefore, the purpose of the present study was to determine whether objective markers of postural dysfunction track disease severity and altered sensory reweighting across the HD spectrum. We hypothesized that low-frequency oscillatory power in the COP signal, and its modulation when vision is removed, would scale with disease stage and dissociate manifest HD from premanifest carriers and family controls. Identifying such signatures could provide objective markers of disease conversion and inform targeted interventions before overt motor diagnosis.

## Methods

### Participants

This cross-sectional observational study enrolled 97 adults (46 ± 14 years, 55 females; Table 1). Participants were divided into four groups: 29 with a diagnosis of Huntington’s disease, 27 premanifest (PM), 28 family members who were not genetically at risk (AR-), and 13 age-matched healthy controls (HC). HC participants provided normative comparison data, whereas AR- participants served as familial/environmental controls. Participants were recruited from the Huntington’s Disease Society of America Center of Excellence at UCSD. This study was approved by the UCSD Institutional Review Board and conducted in accordance with the Code of Federal Regulations for the Protection of Human Subjects.

**Table 1.**
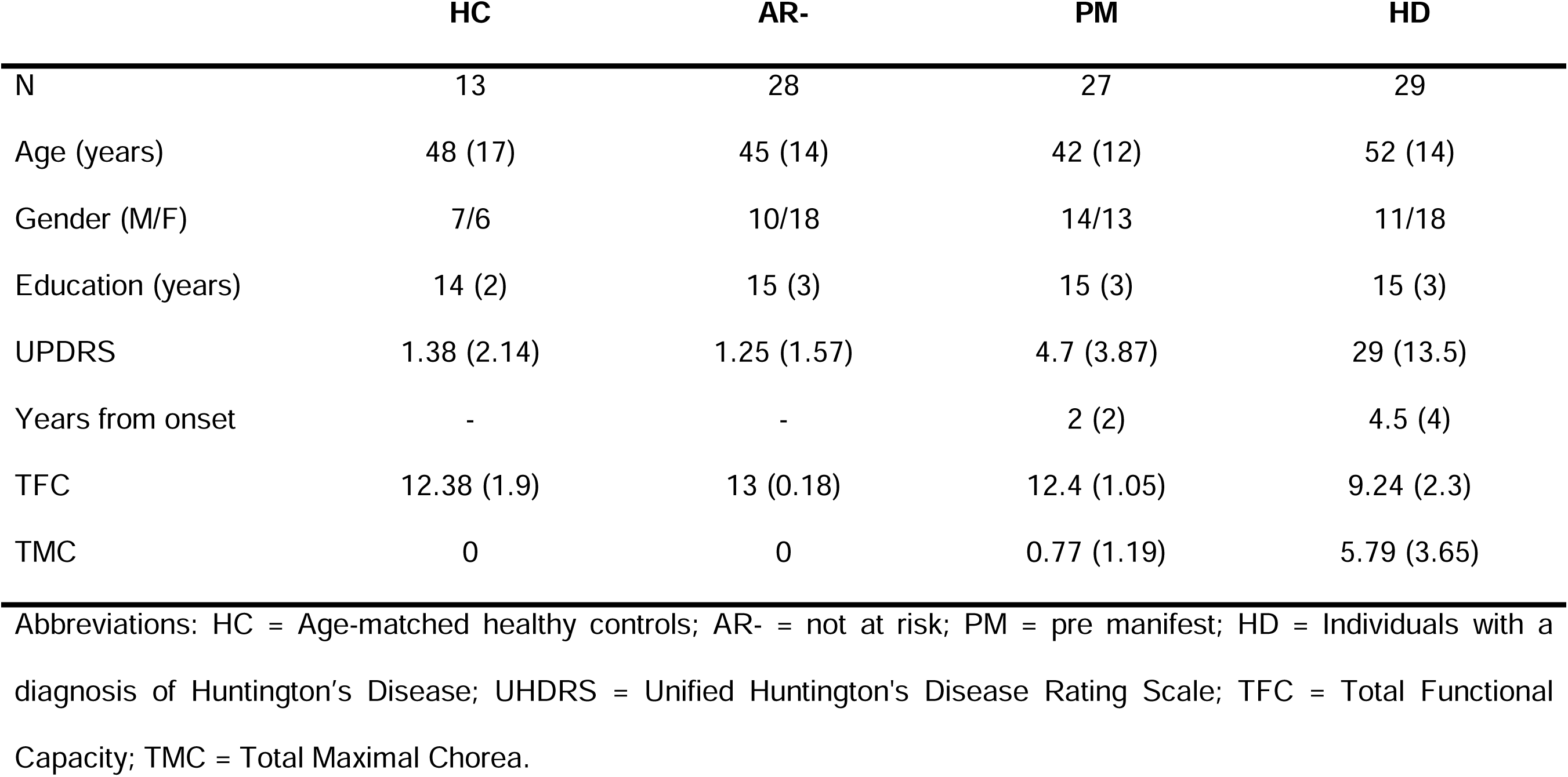
Demographics and clinical characteristics with sample means and standard deviations.

|  | HC | AR- | PM | HD |
| --- | --- | --- | --- | --- |
| N | 13 | 28 | 27 | 29 |
| Age (years) | 48 (17) | 45 (14) | 42 (12) | 52 (14) |
| Gender (M/F) | 7/6 | 10/18 | 14/13 | 11/18 |
| Education (years) | 14 (2) | 15 (3) | 15 (3) | 15 (3) |
| UPDRS | 1.38 (2.14) | 1.25 (1.57) | 4.7 (3.87) | 29 (13.5) |
| Years from onset | - | - | 2 (2) | 4.5 (4) |
| TFC | 12.38 (1.9) | 13 (0.18) | 12.4 (1.05) | 9.24 (2.3) |
| TMC | 0 | 0 | 0.77 (1.19) | 5.79 (3.65) |

Abbreviations: HC = Age-matched healthy controls; AR- = not at risk; PM = pre manifest; HD = Individuals with a diagnosis of Huntington's Disease; UHDRS = Unified Huntington's Disease Rating Scale; TFC = Total Functional Capacity; TMC = Total Maximal Chorea.

Inclusion criteria for HD and PM participants included confirmed CAG repeat length ≥40 and the ability to understand and respond to verbal and written instructions. Exclusion criteria: Individuals with concurrent neurological, cardiovascular, metabolic, or vestibular conditions, as well as those who have recently consumed drugs or alcohol. Individuals were categorized as HD or PM according to Unified Huntington’s Disease Rating Scale (UHDRS)-Total Motor Score (UHDRS-TMS) Diagnostic Confidence Level criteria.^23^

### Equipment

Postural sway was assessed using a force plate and the BTrackS™ Assess Balance System (Balance Tracking Systems, San Diego, CA). Previous studies established the reliability of this system for acquiring COP data related to postural sway.^24–28^ COP data was sampled at 25 Hz and stored on a local PC for offline analysis.

### Experimental Procedures

Each participant completed a single ∼1-hour session, beginning with explanation, demonstration, and familiarization with the equipment and protocol. Participants performed two balance tasks under different vision conditions (Fig. 1):

1. Eyes-open (EO): Participants stood as still as possible on the force plate, feet shoulder-width apart, hands on their hips, for three 10-second trials.
2. Eyes-closed (EC): Participants maintained the same position on the force plate with their eyes closed, across three 10-second trials.

**Fig 1.**
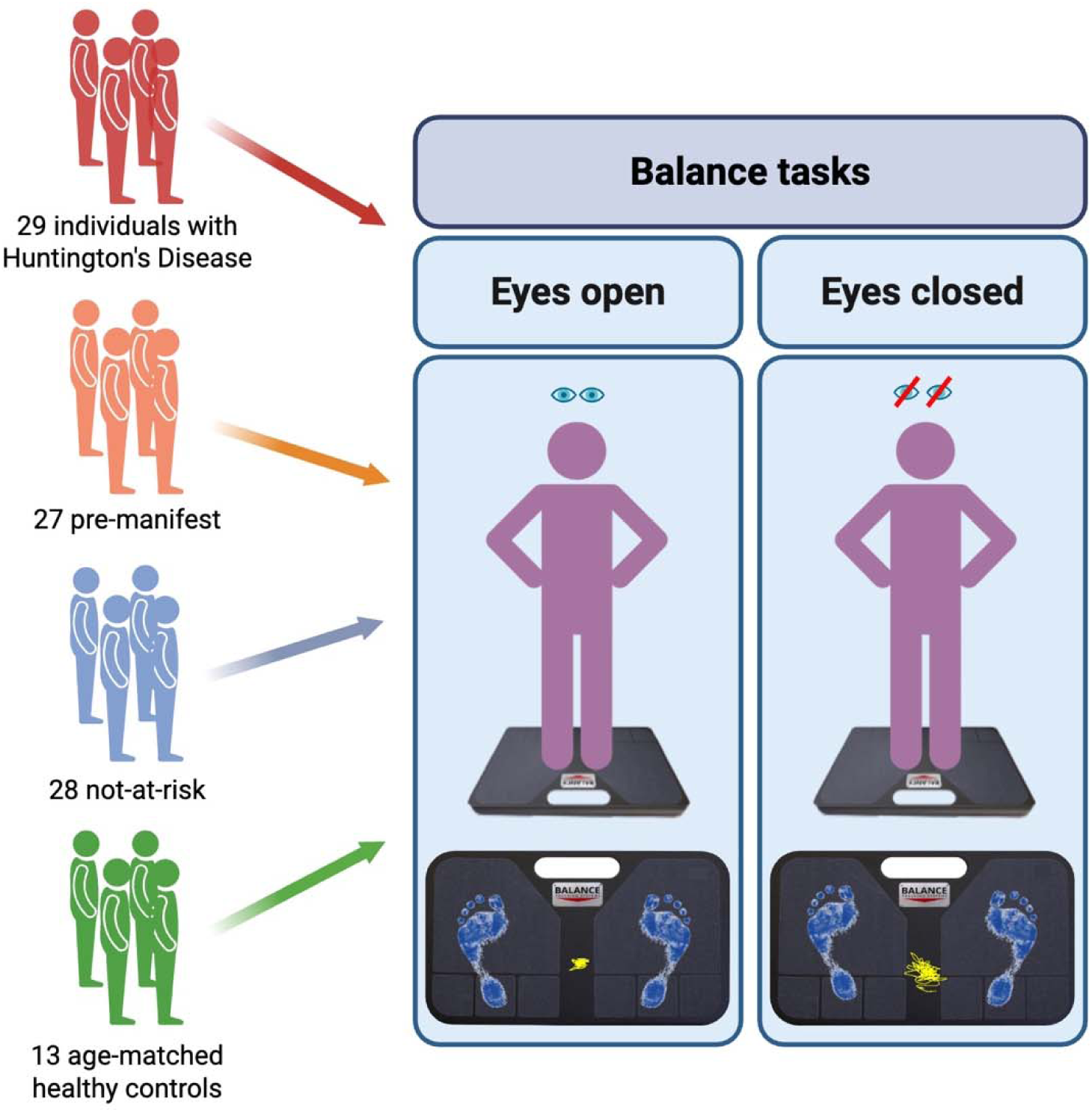
Study design. Ninety-seven adults participated: 29 individuals with Huntington’s Disease, 27 pre-manifest individuals, 28 individuals not at risk, and 13 age-matched healthy controls. All groups completed two quiet standing conditions with eyes open and eyes closed.

Auditory tones marked task onset and offset; task order was randomized and blocked across trials.

### Data Analysis

Data were acquired using the BTrackS Assess Balance software (Version 4, San Diego, CA, USA) and analyzed offline with a custom-written MATLAB (MathWorks Inc., Natick, MA, USA) program. COP signals from each 10-second trial were filtered using a 4th-order Butterworth filter with a low-pass cut-off of 4 Hz. The dependent variables were the anterior-posterior (AP), medial-lateral (ML), and total COP sway displacement (cm). ML and AP sway displacement were calculated using the following formulas:^29^

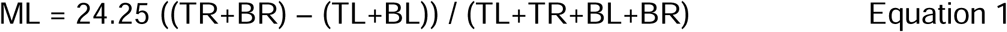

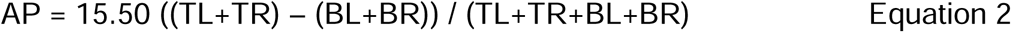

Where TR, TL, BR, and BL are the force sensor values from the top right, top left, bottom right, and bottom left corners of the force plate. The constant 24.25 represents half of the ML width of the force plate, while 15.50 represents half of the AP length of the force plate in cm. The total sway displacement is determined by the distance between the successive registered COP locations according to the following formula:

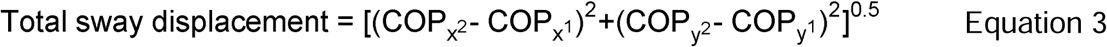

Where COP_x2_ and COP_x1_ are adjacent time points in the COP_x_ (ML) time-series, and COP_y2_ and COP_y1_ are adjacent time points in the COP_y_ (AP) time-series. The total COP sway displacement is then obtained by summing these individual distances across the entire time series ^25^.

In addition, continuous wavelet transform analysis was performed on the COP signal using a base MATLAB function developed by Torrence and Compo (available at: http://paos.colorado.edu/research/wavelets).^30^ For more details about the continuous wavelet transform analysis, refer to Baweja et al. (2011).^31^ For statistical comparisons, the COP signal frequency data were divided into three frequency bands: 0–1 Hz, 1–2 Hz, and 2–4 Hz. Frequency bands were selected based on prior work suggesting that physiologically relevant COP oscillations primarily occur below 4 Hz. The dependent variable was absolute peak power in each band.

### Statistical Analysis

A two-way mixed ANOVA (4 groups × 2 vision conditions),with repeated measures on vision condition and frequency band, compared absolute wavelet power from 0–4 Hz. A three-way mixed ANOVA (4 groups × 2 vision conditions × 3 frequency bands), with repeated measures on vision condition and frequency band, compared the absolute wavelet power from 0-4 Hz.

Multiple linear regression (stepwise) was used to determine the association between the change in wavelet power in the 0-1 Hz band from eyes-open to eyes-closed conditions and the change in total COP sway displacement. This change was calculated using the following formula:^32^

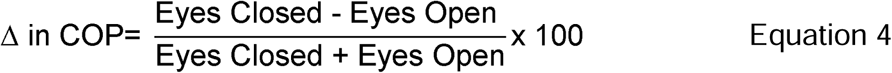

The relative importance of the predictors was estimated with the part correlations (part r), which provide the correlation between a predictor and the criterion. The unique contribution of the change in wavelet power from 0-1 Hz to the change in the total COP sway displacement from eyes-open to eyes-closed was used to examine this relation.

All analyses were performed with the IBM SPSS Statistics v29 package (IBM^®^ SPSS^®^ Inc., Chicago, IL, USA). No participants exceeded the predefined outlier threshold of ±3 SD. Appropriate post hoc analyses were conducted to follow up on significant interactions from the ANOVA models. The alpha level for all statistical tests was 0.05 (unless corrected). Data are reported as mean ± SD within the text and as mean ± SEM in the figures. Only statistically significant findings are discussed in detail.

## Results

### Postural sway

#### Anterior-posterior sway displacement

HD exhibited greater AP sway displacement (*F*_3,93_ = 12.949, *P* < 0.001, *η_p_*^2^ = 0.295) than PM, AR-, and HC groups. The task main effect was significant (*F*_1,93_ = 28.362, *P* < 0.001, *η_p_*^2^ = 0.234), demonstrating greater AP sway displacement with eyes closed than eyes open. The group × task interaction was also significant (*F*_3,93_ = 6.269, *P* < 0.001, *η_p_*^2^ = 0.168; Fig. 2A). HD and PM exhibited greater AP sway displacement with eyes closed than open.

**Fig. 2.**
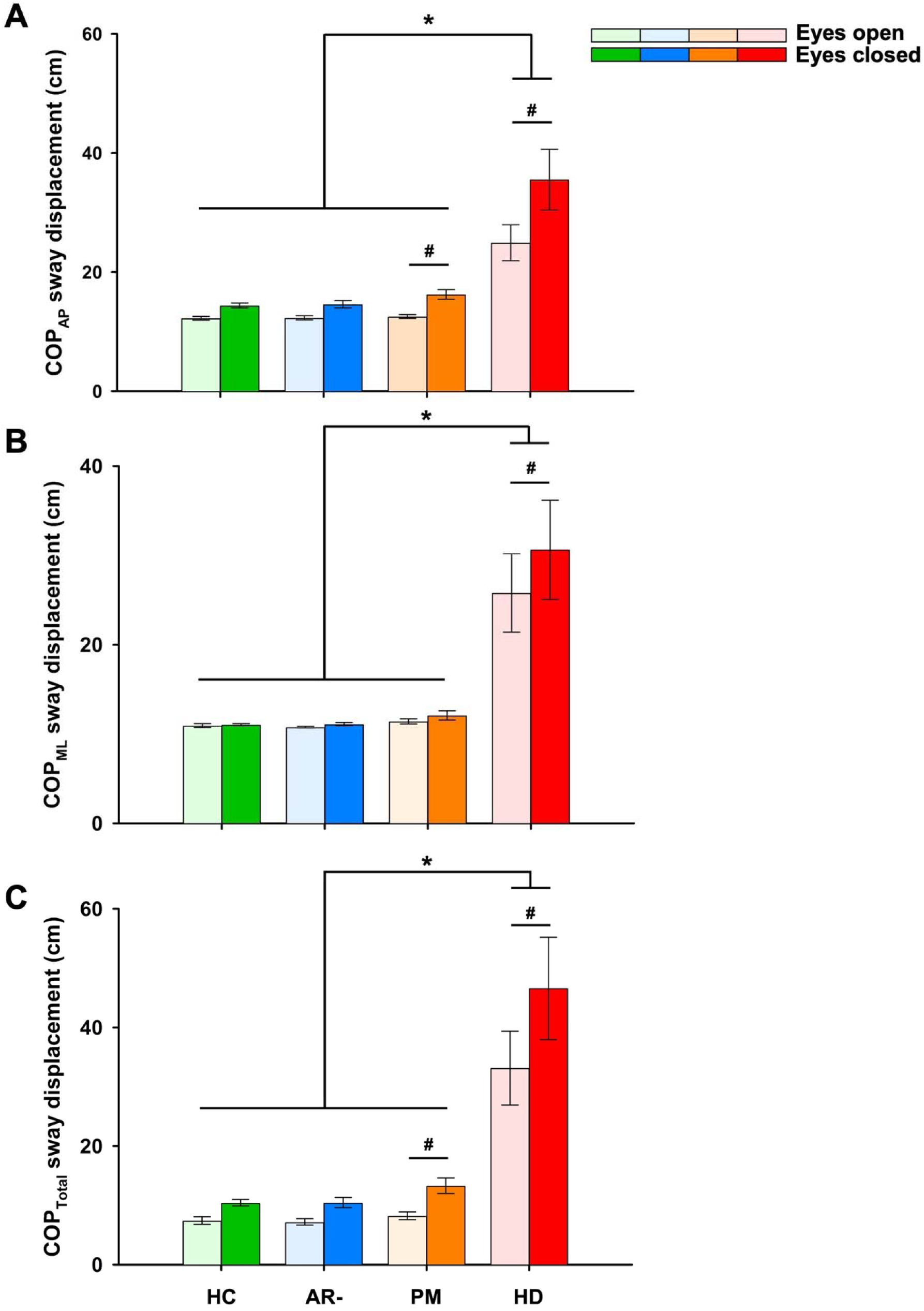
Average postural sway metrics. HD exhibited greater **(A)** AP, **(B)** ML, and **(C)** total COP sway when compared with all other groups. Eyes-closed increase AP and total COP sway in HD and PM groups. *Group. # Task.

#### Medial-lateral sway displacement

HD exhibited greater ML sway displacement (*F*_3,93_ = 9.287, *P* < 0.001, *η_p_*^2^ = 0.231) than the other groups. The task main effect was significant (*F*_1,93_ = 4.407, *P* = 0.038, *η_p_* ^2^ = 0.045), highlighting greater ML sway displacement with eyes closed than open. The group × task interaction was also significant (*F*_3,93_ = 3.066, *P* = 0.032, *η_p_*^2^ = 0.090; Fig. 2B). HD exhibited greater ML sway with eyes closed than open.

#### Total COP sway displacement

HD exhibited greater total COP sway displacement (*F*_3,93_ = 12.996, *P* < 0.001, *η_p_*^2^ = 0.295) than the other groups. The task main effect was significant (*F*_1,93_ = 28.244, *P* < 0.001, *η_p_*^2^ = 0.233), indicating greater total COP sway displacement with eyes closed than open. The group × task interaction was also significant (*F*_3,93_ = 5.339, *P* = 0.002, *η_p_*^2^ = 0.147; Fig. 2C). The HD and PM groups exhibited greater total COP sway displacement in the eyes closed condition when compared with the eyes open condition.

### Wavelet power spectrum of the COP signal from 0-4 Hz

#### Anterior-posterior 0-4 Hz: absolute wavelet power

HD exhibited greater AP wavelet power (*F*_3,93_ = 13.155, *P* < 0.001, *η_p_*^2^ = 0.298; Fig. 3) when compared with the PM, AR-, and HC groups. The task main effect was significant (*F*_1,93_ = 7.839, *P* = 0.006, *η_p_*^2^ = 0.078), demonstrating greater AP wavelet power with eyes closed than with eyes open. The frequency main effect was significant (*F*_2,186_ = 41.134, *P* < 0.001, *η_p_*^2^ = 0.307), pointing to greater power in the 0-1 Hz band than with 1-2 and 2-4 Hz. Additionally, wavelet power in the 1-2 Hz band was greater when compared with the 2-4 Hz band. The group × task interaction was also significant (*F*_3,93_ = 3.227, *P* = 0.026, *η_p_*^2^ = 0.094). HD exhibited greater AP wavelet in the open and eyes-closed conditions relative to all the other groups. The group × frequency interaction was also significant (*F*_6,186_ = 8.032, *P* < 0.001, *η_p_*^2^ = 0.206). HD exhibited greater AP wavelet power across all frequency bands as compared to all other groups. The task × frequency interaction was significant (*F*_2,186_ = 3.515, *P* = 0.032, *η_p_*^2^ = 0.036), indicating greater AP wavelet power in the eyes closed compared to eyes open condition across all frequency bands. All other main effects and interactions were not significant.

**Fig. 3.**
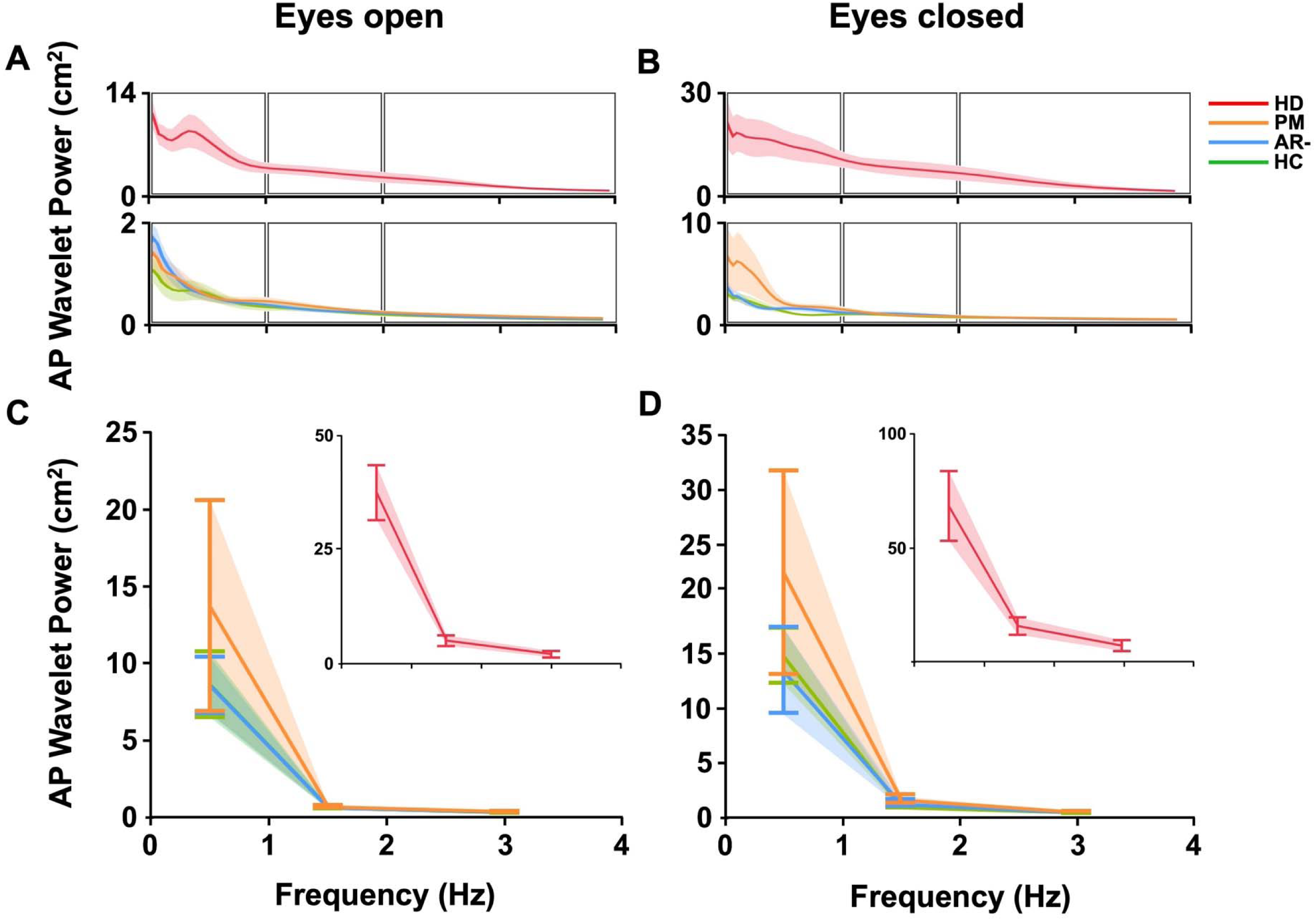
AP wavelet power spectrum (0-4 Hz). **(A-B)** Average power spectrum during eyes open (left) and eyes closed (right) conditions. (**C-D)** HD exhibited greater power across frequencies and conditions. Eyes-closed, increase ML power. 0-1 Hz > 1-2 Hz > 2-4 Hz. *Group. # Task.

#### Medial-lateral 0-4 Hz: absolute wavelet power

The group main effect was significant (*F*_3,93_ = 8.959, *P* < 0.001, *η_p_*^2^ = 0.224; Fig. 4), indicating that HD exhibited greater ML wavelet power compared with the PM, AR-, and HC groups. The frequency main effect was also significant (*F*_2,186_ = 10.642, *P* < 0.001, *η_p_*^2^ = 0.103), demonstrating greater power in the 0-1 Hz band as compared to the 1-2 and 2-4 Hz bands. Additionally, wavelet power in the 1-2 Hz band was greater when compared to the 2-4 Hz band. The group × frequency interaction was also significant (*F*_6,186_ = 7.937, *P* < 0.001, *η_p_*^2^ = 0.204). The HD group exhibited greater ML wavelet power across all frequency bands when compared to the PM, AR-, and HC groups. All other main effects and interactions were not significant.

**Fig. 4.**
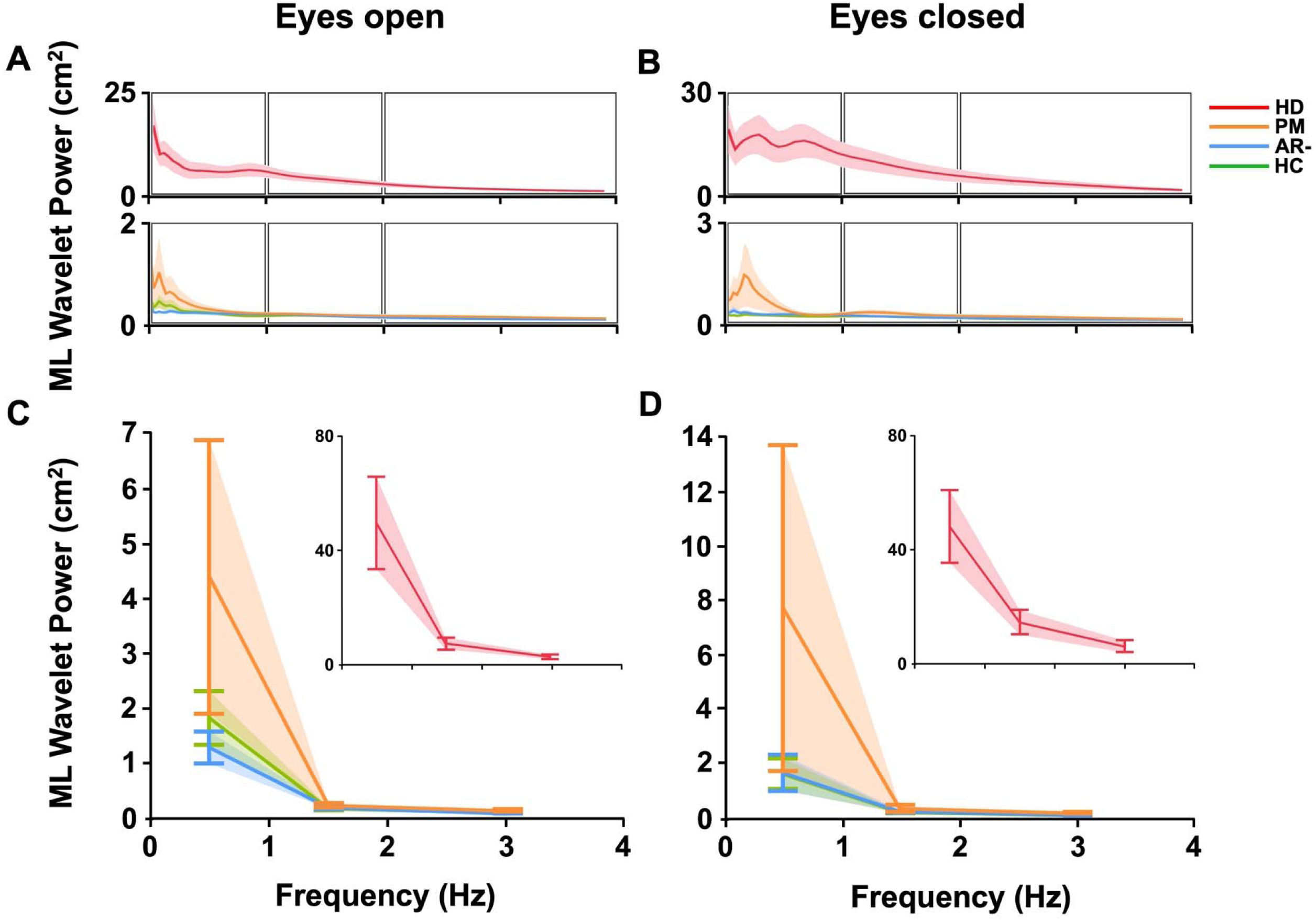
ML wavelet power spectrum (0-4 Hz). **(A-B)** Average power spectrum during eyes-open (left) and eyes-closed (right) conditions. **(C-D)** HD exhibited greater power across frequencies. Power was greatest in 0-1 Hz > 1-2 Hz > 2-4 Hz. *Group. # Task.

#### Total COP 0-4 Hz: absolute wavelet power

HD exhibited greater COP wavelet power when compared to the PM, AR-, and HC groups. The task main effect was significant (*F*_1,93_ = 4.537, *P* = 0.036, *η_p_^2^* = 0.047), signifying greater COP wavelet power with eyes closed than with eyes open. The frequency main effect was significant (*F*_2,186_ = 5.264, *P* = 0.006, *η_p_^2^* = 0.054; Fig. 5), highlighting greater power in the 0-1 Hz band as compared to the 1-4 Hz bands. The group × task interaction was also significant (*F*_3,93_ = 4.714, *P* = 0.004, *η_p_^2^* = 0.132). The HD group exhibited greater COP wavelet power in the eyes-closed condition compared to the eyes-open condition. The group × frequency interaction was also significant (*F*_6,186_ = 5.561, *P* < 0.001, *η_p_^2^* = 0.152). HD exhibited greater COP wavelet power in the 0-1 Hz band compared with the 1-2 and 2-4 Hz bands. The task × frequency interaction was significant (*F*_2,186_ = 3.647, *P* = 0.028, *η_p_^2^* = 0.038). The COP wavelet power in the 0-1 Hz band was greater in the eyes-closed condition compared to the eyes-open condition. The group × task × frequency interaction was significant (*F*_6,186_ = 4.105, *P* < 0.001, *η_p_^2^* = 0.117), demonstrating that during eyes-open and eyes-closed conditions, the HD group exhibited greater COP wavelet power across all frequency bands compared with the PM, AR-, and HC groups.

**Fig. 5.**
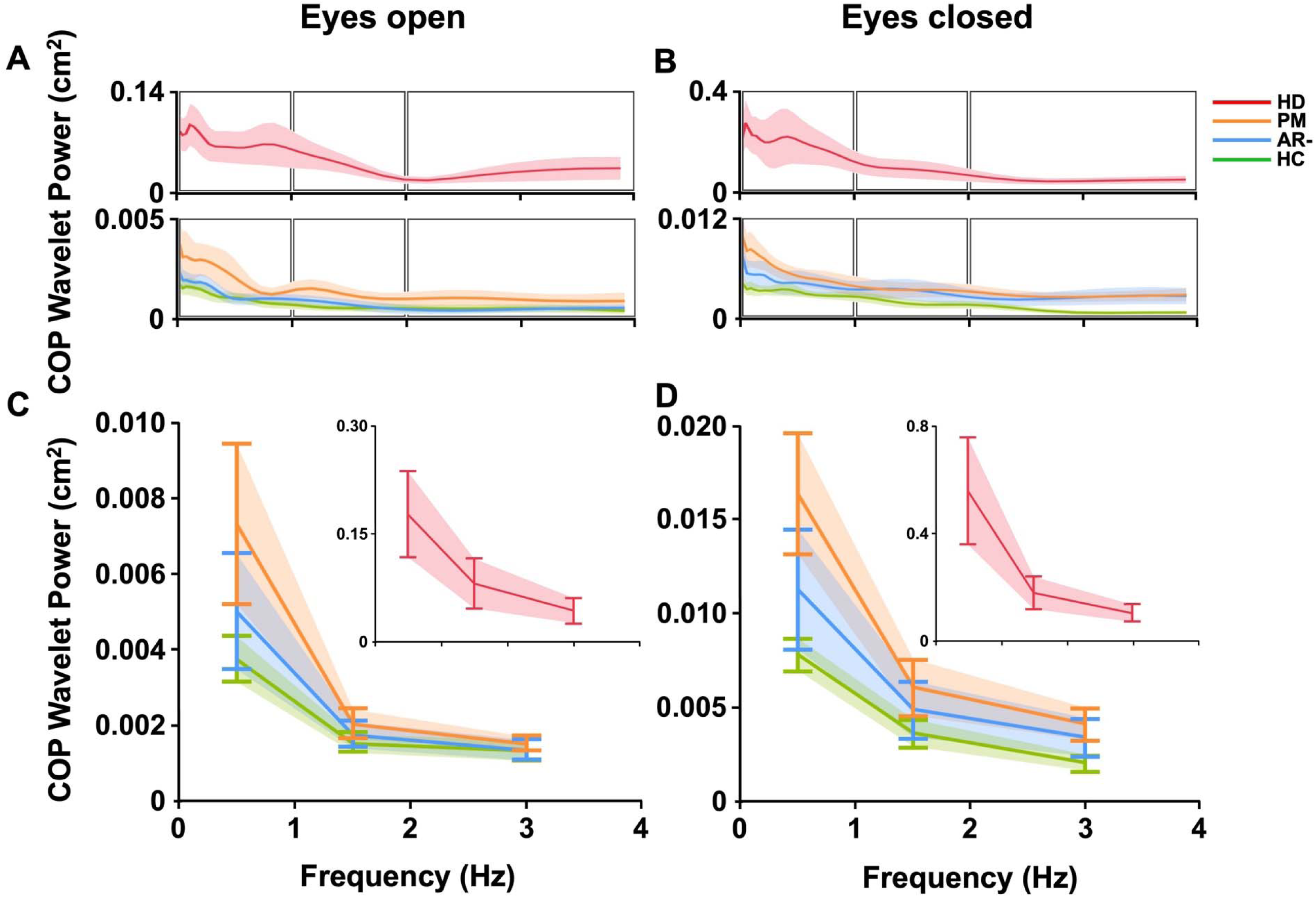
Total COP wavelet power (0-4 Hz). **(A-B)** Average power spectrum during eyes open (left) and eyes closed (right) conditions. **(C-D)** HD exhibited greater power across frequencies and conditions. Eyes closed, increase COP power. 0-1 Hz > 1-2 Hz > 2-4 Hz. *Group. # Task.

### Association between wavelet power from 0-1 Hz and total COP sway displacement

Because both total COP sway displacement and 0-1 Hz wavelet power increased during the eyes-closed condition, we next tested whether the task-related change in low-frequency oscillatory power was functionally coupled to the corresponding change in postural sway. We operationally defined coupling as the extent to which the eyes-open to eyes-closed change in 0-1 Hz COP wavelet power predicted the concurrent change in total COP sway displacement. In this framework, stronger coupling indicates that low-frequency oscillatory modulation closely tracks with the behavioral sway response to sensory challenge, consistent with adaptive sensory reweighting. Conversely, weaker coupling indicates that low-frequency oscillatory activity is present but is less directly translated into organized postural output.

Across the full sample, the eyes-open to eyes-closed change in 0–1 Hz wavelet power significantly predicted the corresponding change in total COP sway displacement, explaining 35% of the variance in sway change (R² = 0.35, P < 0.001), and suggesting that low-frequency oscillatory modulation is closely linked to the magnitude of the postural sway response during sensory challenge. However, the strength of this oscillatory-sway coupling differed substantially across groups. The relationship was strongest in healthy controls, where 0-1 Hz power modulation explained 68% of the variance in total sway change (R^2^ = 0.68, *P* < 0.001; Fig. 6A). Comparable but moderately weaker associations were observed in the AR- and PM groups, where low-frequency power modulation explained 45% and 46% of the variance, respectively (AR-: R^2^ = 0.45, *P* < 0.001; Fig. 6B; PM: R^2^ = 0.46, *P* < 0.001; Fig. 6C). In contrast, this relationship was markedly attenuated in manifest HD, where 0-1 Hz power modulation explained only 14% of the variance in total sway change (R^2^ = 0.14, *P* = 0.041; Fig. 6D). These findings indicate that low-frequency oscillatory activity remains behaviorally coupled to postural sway in HC, AR-, and PM individuals, but is substantially degraded in manifest HD. Thus, although HD individuals exhibited elevated low-frequency power and greater sway, the increase in 0-1 Hz oscillatory activity no longer closely tracked postural sway magnitude.

**Fig. 6.**
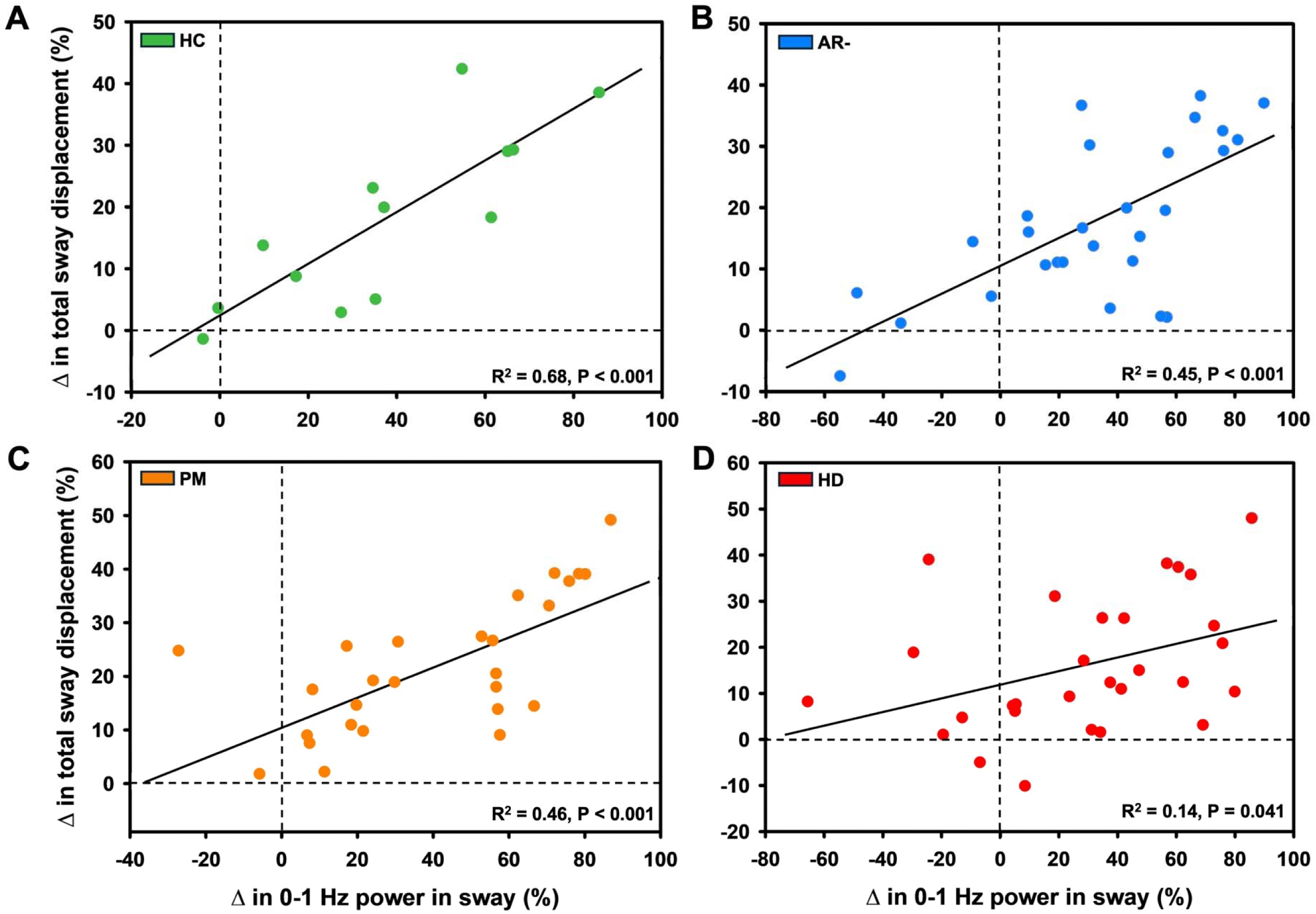
Association between the change in wavelet power in the 0-1 Hz band from eyes-open to eyes-closed and total COP sway displacement. Sensory reweighing at 0-1 Hz accounted for **(A)** 68% of HC, **(B)** 45% of AR-, **(C)** 46% of PM, and **(D)** 14% of HD of COP displacement variance.

## Discussion

The principal finding of this study is that the functional coupling between low-frequency COP oscillations and postural sway progressively weakens across the HD spectrum and becomes markedly degraded in manifest HD. Manifest HD individuals exhibited greater postural sway in every direction compared with the PM, AR-, and HC groups. HD and PM groups, but not AR- or HC, exhibited greater postural sway with eyes closed. Across all groups, most wavelet power between 0 and 4 Hz was concentrated below 1 Hz. The eyes-open to eyes-closed change in 0-1 Hz wavelet power was associated with an increase in total COP sway displacement across all groups, but this relation was substantially weaker in HD individuals (14% variance explained), when compared with HC (68%), AR- (45%), and PM (46%) groups.

### Postural control deficits emerge in premanifest HD and worsen with disease progression

Manifest HD individuals exhibited greater postural sway in every direction (AP, ML, and total COP sway displacement) when compared with the PM, AR-, and HC groups (Fig. 2). These postural control deficits are consistent with disrupted multisensory integration from basal ganglia degeneration, translating into an elevated fall risk.^10,33^

Critically, the HD and PM, but not the AR- or HC, groups showed a significant increase in AP and total COP sway displacement when vision was removed (Fig. 2A-C), aligning with prior reports that postural control impairment begins in the premanifest stage and worsens across the transition to manifest HD.^9-12^ The fact that PM individuals destabilize specifically when vision is removed, yet remain comparable to controls with eyes open, indicates an early deficit in sensory reweighting that manifests only when visual input is unavailable, emerging prior to overt motor diagnosis.

However, linear sway measures alone have been criticized for collapsing the rich temporal structure of the COP signal into a single magnitude and providing limited mechanistic insight.^34,35^ To address this limitation, we examined the time-frequency structure of the COP signal, which has not previously been characterized across the full HD spectrum, and can expose distinct sensory contributions to postural control.

### Frequency signatures distinguish manifest HD from premanifest stages

Across all groups, wavelet power was concentrated in the 0-1 Hz band relative to the 1-2 and 2-4 Hz bands, consistent with prior evidence that the most behaviorally relevant information about postural sway lies below 1 Hz (Fig. 3-5).^16,19,21^ However, manifest HD individuals showed significantly greater wavelet power in every frequency band in both eyes-open and eyes-closed conditions. The elevated low-frequency power even with eyes open indicates that HD individuals are unable to appropriately weight available visual, proprioceptive, and vestibular inputs at baseline. The further increase with eyes closed indicates an additional failure of proprioceptive reweighting once vision is removed.^36^ This is mechanistically consistent with basal ganglia-dependent disruption of proprioceptive integration, a pattern also reported in other basal ganglia disorders, such as Parkinson’s disease.^37–39^

### Oscillatory–sway coupling is preserved in PM but degraded in manifest HD

The key mechanistic finding from the regression analysis is that disease progression in HD appears to weaken the functional coupling between low-frequency COP oscillations and postural sway (Fig. 6). Disease progression in HD is associated not simply with more sway or greater low-frequency power, but with a breakdown in the functional relationship between the two. In healthy postural control, low-frequency COP oscillations below 1 Hz are thought to reflect slow, sensory-guided corrections that help maintain upright stance as sensory input changes.^22,40^ When vision is removed, the nervous system must increase reliance on proprioceptive and vestibular information. In this context, a proportional increase in 0-1 Hz power that explains the increase in total sway can be interpreted as preserved oscillatory-sway coupling -- the time-frequency structure of the COP signal remains organized relative to the behavioral postural response.

This coupling was preserved in HC, AR-, and PM individuals, suggesting that low-frequency modulation remained meaningfully linked to postural output even in the premanifest stage. The PM group showed greater sway when vision was removed, but the regression pattern indicates that their 0-1 Hz oscillatory response still scaled with the resulting sway (Fig. 6C). This may reflect an early sensory reweighting deficit, but not a complete failure of the postural control system to translate low-frequency sensory corrections into behavioral output. The finding that AR- individuals performed similarly to healthy controls is important because it strengthens the disease-specific interpretation of these results. Since AR- individuals share familial and environmental contexts with the HD cohort but do not carry the pathogenic expansion, their preserved postural control suggests that the deficits observed in PM and HD are more likely attributable to HD-related pathophysiology rather than shared family background or environmental exposure.

In contrast, manifest HD showed clear oscillatory-sway decoupling. Although the HD group exhibited greater 0-1 Hz power and greater total sway, the change in low-frequency power explained only a small proportion of the change in sway (Fig. 6D). This suggests that low-frequency oscillations are still being generated, but are no longer effectively coupled to organized postural adjustments. Mechanistically, this pattern is consistent with impaired basal ganglia-mediated integration of visual, vestibular, and proprioceptive inputs.^5,6^ Rather than reflecting adaptive sensory reweighting, elevated low-frequency power in manifest HD may reflect poorly organized corrective activity, noisy sensorimotor output, or compensatory oscillations that fail to stabilize the body effectively. Therefore, the transition from PM to manifest HD may be marked by a shift from preserved but challenged sensory reweighting to a decoupled postural control state in which oscillatory activity becomes progressively less functional.

In conclusion, postural control deficits progress across the HD spectrum, culminating in increased sway, elevated low-frequency COP power, and weakened sensory reweighting in manifest HD. PM individuals show early balance vulnerability when vision is removed, but retain relatively preserved low-frequency COP structure. Across groups, COP power was concentrated below 1 Hz; however, 0-1 Hz oscillatory-sway coupling was preserved in HC, AR-, and PM individuals but weakened in manifest HD. This stage-dependent decoupling may mark the transition from compensatory sensory reweighting to impaired postural control.

## Author Roles

M.M.V.: Statistical analysis: A. Design, B. Execution; Manuscript preparation: A. Writing of the first draft

T.W.: Statistical analysis: Execution; Manuscript preparation: A. Writing of the first draft,

K.T.G.: Research project: Organization, Execution Statistical analysis: A. Design, B. Execution; Manuscript preparation: A. Writing of the first draft

A.J.L.: Research project: Organization, Execution Statistical analysis: A. Design, B. Execution; Manuscript preparation: A. Writing of the first draft

J.P.M.: Research project: Organization, Execution Statistical analysis: A. Design, B. Execution; Manuscript preparation: A. Writing of the first draft

A.J.P.: Research project: Organization, Execution Statistical analysis: A. Design, B. Execution; Manuscript preparation: A. Writing of the first draft

N.B.: Conception, Organization; Manuscript preparation: Review and critique

D.J.G.: Conception, Organization, Execution; Manuscript preparation: Review and critique

P.E.G.: Conception, Organization, Execution; Manuscript preparation: Review and critique

J.C.-B.: Execution, Review and critique, Manuscript preparation

H.S.B.: Conception, Organization, Execution; Statistical analysis: A. Design, B. Execution, C. Review and critique; Manuscript preparation: A. Writing of the first draft, B. Review and critique

## Data Availability

All data produced in the present study are available upon reasonable request to the authors

## Acknowledgments

The authors have no acknowledgments to report.

## Disclosures

### Ethical Compliance Statement

Institutional Review Board approval was obtained from the University of California, San Diego Institutional Review Board (protocol no. 170038). Written informed consent was obtained from all participants before participation. We confirm that we have read the Journal’s position on issues involved in ethical publication and affirm that this work is consistent with those guidelines.

### Financial Disclosures for the Previous 12 Months

DJ. Goble is eligible for royalties from a patent (US Patent 10,660,558,2020) related to the BTrackS Balance Plate. In addition, DJG has an equity stake (stock options) in Balance Tracking Systems, Inc. This financial conflict of interest is mitigated by a management plan implemented by his academic institution to ensure the integrity of research.

